# Determinants of Persistent Post COVID-19 symptoms: Value of a Novel COVID-19 symptoms score

**DOI:** 10.1101/2020.11.11.20230052

**Authors:** Islam Galal, Aliae AR Mohamed Hussein, Mariam T Amin, Mahmoud M Saad, Hossam Eldeen E Zayan, Mustafa Z Abdelsayed, Mohamed M Moustafa, Abdel Rahman Ezzat, Radwa ED Helmy, Howaida K Abd- Elaal, Nasrallah A Al Massry, Mohamed A. Soliman, Asmaa M Ismail, Karima MS Kholief, Enas Fathy, Maiada K Hashem

## Abstract

**Background:** Being a newly emerging disease little is known about its long-lasting post COVID-19 consequences. Aim of this work is to assess the frequency, patterns and determinants of persistent post COVID-19 symptoms and to evaluate the value of a proposed Novel COVID-19 symptoms score. Patients with confirmed COVID-19 in the registry were included in a cross sectional study. The patient demographics, comorbid disorders, the mean duration since the onset of the symptoms, history of hospital or ICU admittance, and treatment taken during acute state, as well as symptoms score before and after convalescence were recorded.

**Results:** The most frequent constitutional and neurological symptoms were myalgia (60.0%), arthralgia (57.2%), restriction of daily activities (57.0%), sleeping troubles (50.9%), followed by anorexia (42.6%), chest pain (32.6%), gastritis (32.3%), cough (29.3%) and dyspnea (29.1%). The mean total score of acute stage symptoms was 31.0 ± 16.3 while post COVID 19 symptoms score was 13.1±12.6 (P<0.001). The main determinants of the persistent post COVID-19 symptoms were the need for oxygen therapy (P<0.001), pre-existing hypertension (P=0.039), chronic pulmonary disorders (P=0.012), and any chronic comorbidity (P=0.004). There was a correlation between the symptom score during the acute attack and post COVID-19 stage (P<0.001, r=0.67). The acute phase score had 83.5% sensitivity and 73.3% specificity for the cutoff point > 18 to predict occurrence of Post-COVID-19 symptoms.

**Conclusions:** COVID-19 can present with a diverse spectrum of long-term post COVID-19 symptoms. Increased acute phase symptom severity and COVID-19 symptom score > 18 together with the presence of any comorbid diseases increase the risk for persistent post COVID-19 manifestations and severity.

## Background

Being a newly emerging disease a little known about long-lasting post COVID-19 infection consequences. However, in the coming days, great stress will progressively comprise post-acute carefulness of those recovered cases from COVID-19. It is expected that COVID-19 may have a principal effect on the physical, mental, cognitive, and public health state(1). Moreover, numerous serious COVID-19 infections necessitate intensive care unit (ICU) management and may lead to persistent post-convalescence consequences comprising respiratory, somatic, mental, and emotional abnormalities which known as post-intensive care syndrome (2-4). Recent studies illustrated that in patients who had convalesced from COVID-19, about 50-87% experienced persistence of at least one symptom, predominantly lethargy and shortness of breath that may necessitate some form of constant carefulness to recover their long-lasting consequences (5) (6). High blood pressure, obesity, and mental health conditions are proposed risk factors for persistent post COVID symptoms, however, surprisingly, even young adults and children without underlying chronic medical conditions and those with mild COVID-19 illness reported that they had not returned to their usual state of health several weeks after convalescence(7). So, further researches are needed to understand the risk factors and pathophysiology of these persistent post COVID-19 symptoms.

### Aim of this work is to

Assess the frequency, patterns and determinants of persistent post COVID-19 symptoms and to evaluate the value of a proposed Novel COVID-19 symptoms score.

## Methods

The present study was a cross-sectional study performed during 18^th^ of July to 31 of August 2020. Patients were included if they had confirmed COVID-19 in the registry (positive or indeterminate COVID-19 PCR test, or presumed presence of Covid-19 based on clinical and radiological criteria). They were interviewed in the follow-up clinics and filled paper follow-up forms. Medical students, residents and volunteers evaluated patient’s symptoms and revised the submitted forms for missing data.

### Sample size

was calculated using Epi info statistical package version 7. Based on number of cases in Egypt on 18^th^ July 26, 2020 (86474), the following parameters for cross-sectional study were expected cases with 0.50, with acceptable margin of error 0.05, design effect 1, 95% confidence level. The required sample size was 384 patients. It was raised to 425 after considering 10% as a drop out.

**The following data were collected:**

- The patient demographics including age, gender, body mass index, smoking status, history of comorbid disorders, the mean duration since the onset of the symptoms, history of hospital or ICU admittance, and treatment taken during the acute attack were recorded
- Symptoms during the acute attack of COVID-19.
- Symptoms after the convalescence from the acute attack of COVID-19 including general, upper, lower respiratory tract, neurological, cutaneous complains and symptoms suggesting other system of the body affection.

### Symptoms Score

Two scores were used. Acute stage symptoms include 27 symptoms and post-COVID symptoms include 29 symptoms. A 4-point Likert scale was used for each symptom reported as absent, mild, moderate, or severe. For acute stage and post-COVID symptoms, the range of the overall score is 0–81 and 0 – 87 respectively (the higher the number, the further symptom severity).

**The study was approved by** the ethical committee of Aswan Faculty of Medicine, Egypt **(IRB number: aswu /469/7/2020)** and registered in Clinicaltrial.gov: NCT04479293.

### Statistical Analysis

Statistical analyses were performed using IBM SPSS Statistics version 20 (SPSS Inc., Chicago, IL, USA). Categorical data were presented as numbers and percentages, while continuous data were reported as means ± SD and/or Median (min -max) and tested for normality using the Shapiro-Wilkes test. As symptom score was not normally distributed the Mann-Whitney and Kruskal-Wallis tests were used to compare score according to different variables. Also, Spearman’s correlation was used to find the correlation between symptoms’ score in acute and post COVID-19 stages. TO detect the sensitivity and accuracy of acute stage symptoms’ score in prediction of persistent Post-COVID symptoms, ROC curve analysis was performed. In all statistical tests, p-value <0.05 was considered statistically significant.

## Results

The study involved 430 participants. They were 156 males and 274 females, their mean age was 37.4 ± 12.6 years, and the range was (12-74 years). The most common presenting symptoms during acute attack were myalgia (89.5%), fever (85.1%), restriction of daily activities (84.9%), while the least frequent complaint was memory loss (21.2%). Among them, 370 (86%) reported persistent post COVID-19 symptoms. The most frequent constitutional and neurological symptoms were myalgia (60.0%), arthralgia (57.2%), restriction of daily activities (57.0%), sleeping troubles (50.9%), nervousness and hopelessness (53.3%), while the most common respiratory and GIT symptoms were anorexia (42.6%), chest pain (32.6%), gastritis (32.3%), cough (29.3%) and dyspnea (29.1%), while the least frequent symptom was newly diagnosed DM (8.8%) ***(Figure 1 a-c)***. The mean total score of acute stage symptoms was 31.0 ± 16.3 while post COVID 19 symptoms score was 13.1±12.6 (P<0.001).

**Figure 1.**
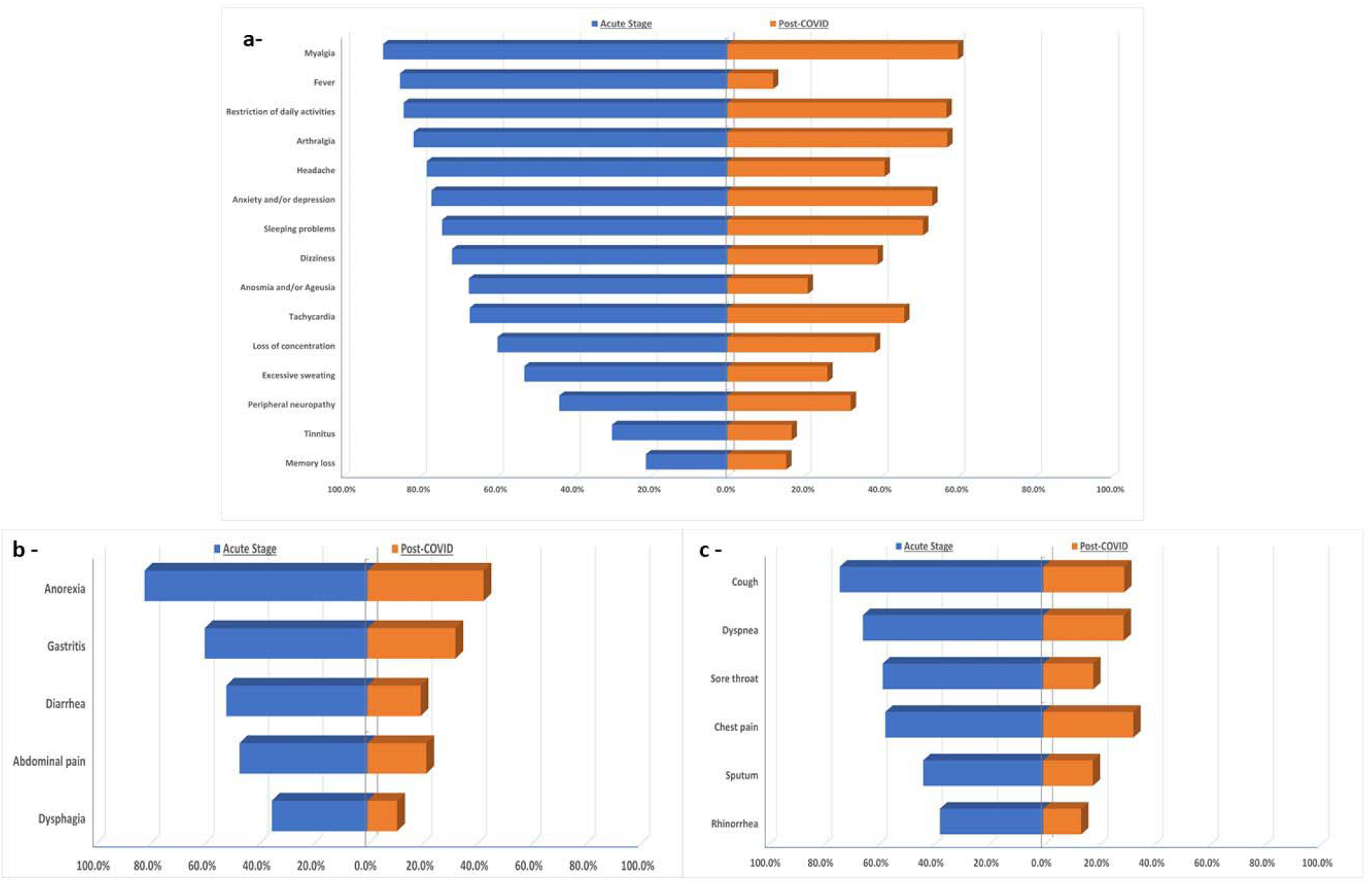
Reported symptoms in acute and post-COVID stages: a-General symptoms, b-GIT symptoms, c-Respiratory symptoms.

The main essential determinant of the persistent post COVID-19 symptoms and symptom score among the included patients was the need for oxygen therapy (P< 0.001) as shown in ***Table (1)***.

**Table 1.** Determinants of persistent post COVID-19 symptoms and symptom score in the included patients (n=430)

|  | N (%) | Mean Score | P-value * |
| --- | --- | --- | --- |
| Age (years) |  |  |  |
| < 25 | 58 (13.5%) | 12.9 ± 13.5 | 0.393 |
| 25 – 40 | 227 (52.8%) | 12.6 ± 12.3 |  |
| > 40 | 145 (33.7%) | 14.0 ± 12.7 |  |
| Gender |  |  |  |
| Male | 156 (36.3%) | 13.2 ± 12.5 | 0.998 |
| Female | 274 (63.7%) | 13.1 ± 12.6 |  |
| BMI |  |  |  |
| Underweight | 147 (34.2%) | 11.9 ± 13.6 | 0.107 |
| Normal | 120 (27.9%) | 13.6 ± 11.4 |  |
| Overweight | 52 (12.1%) | 14.4 ± 15.2 |  |
| Obese | 66 (15.3%) | 14.5 ± 12.6 |  |
| Smoking |  |  |  |
| Nonsmoker | 371 (86.3%) | 12.9 ± 12.8 | 0.138 |
| Current smoker | 26 (6%) | 13.3 ± 10.8 |  |
| Ex-smoker | 33 (7.7%) | 16.1 ± 11.8 |  |
| Time elapsed since COVID-19 diagnosis |  |  |  |
| ≤ One month | 194 (45.1%) | 13.5 ± 11.5 | 0.132 |
| > One month | 236 (54.9%) | 12.8 ± 13.5 |  |
| Hospital admission during illness |  |  |  |
| Yes | 103 (24%) | 14.0 ± 12.4 | 0.216 |
| No | 327 (76%) | 12.8 ± 12.7 |  |
| Need of oxygen therapy |  |  |  |
| Yes | 72 (16.7%) | 17.4 ± 12.5 | < 0.001 ^ |
| No | 358 (83.3%) | 12.3 ± 12.4 |  |
| ICU admission |  |  |  |
| Yes | 20 (4.7%) | 17.3 ± 12.3 | 0.066 |
| No | 410 (95.3%) | 12.9 ± 12.6 |  |

\* Mann-Whitney & Kruskal-Wallis tests were used
^ significant p-value.

There were 26.5% patients reported that they have a chronic illness and distribution of those conditions illustrated in ***figure (2)***. The most frequent pre-existing co-morbidities allied with the persistent post COVID-19 symptoms and symptom score among the study population were hypertension (P= 0.039) followed by chronic pulmonary disorders (P= 0.012), and lastly the presence of any chronic disorder (P= 0.004), as shown in ***Table (2)***. There was no difference in post COVID-19 symptoms and symptom score despite difference in received treatment (supportive treatment, hydroxychloroquine, azithromycin or corticosteroids).

**Table 2.** Post COVID-19 symptoms’ score in patients according to comorbidities (n=430)

|  | Yes | No | P-value * |
| --- | --- | --- | --- |
| **Diabetes Mellitus** | 13.4 ± 11.8 | 13.1 ± 12.7 | 0.730 |
| **Hypertension** | 15.4 ± 12.9 | 12.7 ± 12.5 | 0.039 ^ |
| **Cardiac disease** | 13.8 ± 4.9 | 13.1 ± 12.7 | 0.276 |
| **Chronic pulmonary disease** | 20.5 ± 15.6 | 12.7 ± 12.3 | 0.012 ^ |
| **Renal disease** | 16.5 ± 11.9 | 13.1 ± 12.6 | 0.278 |
| **Psychiatric disease** | 22.0 ± 14.2 | 12.9 ± 12.5 | 0.068 |
| **Any chronic illness** | 15.8 ± 13.6 | 12.2 ± 12.1 | 0.004 ^ |

\* Mann-Whitney test were uses
^ significant p-value.

**Figure 2.**
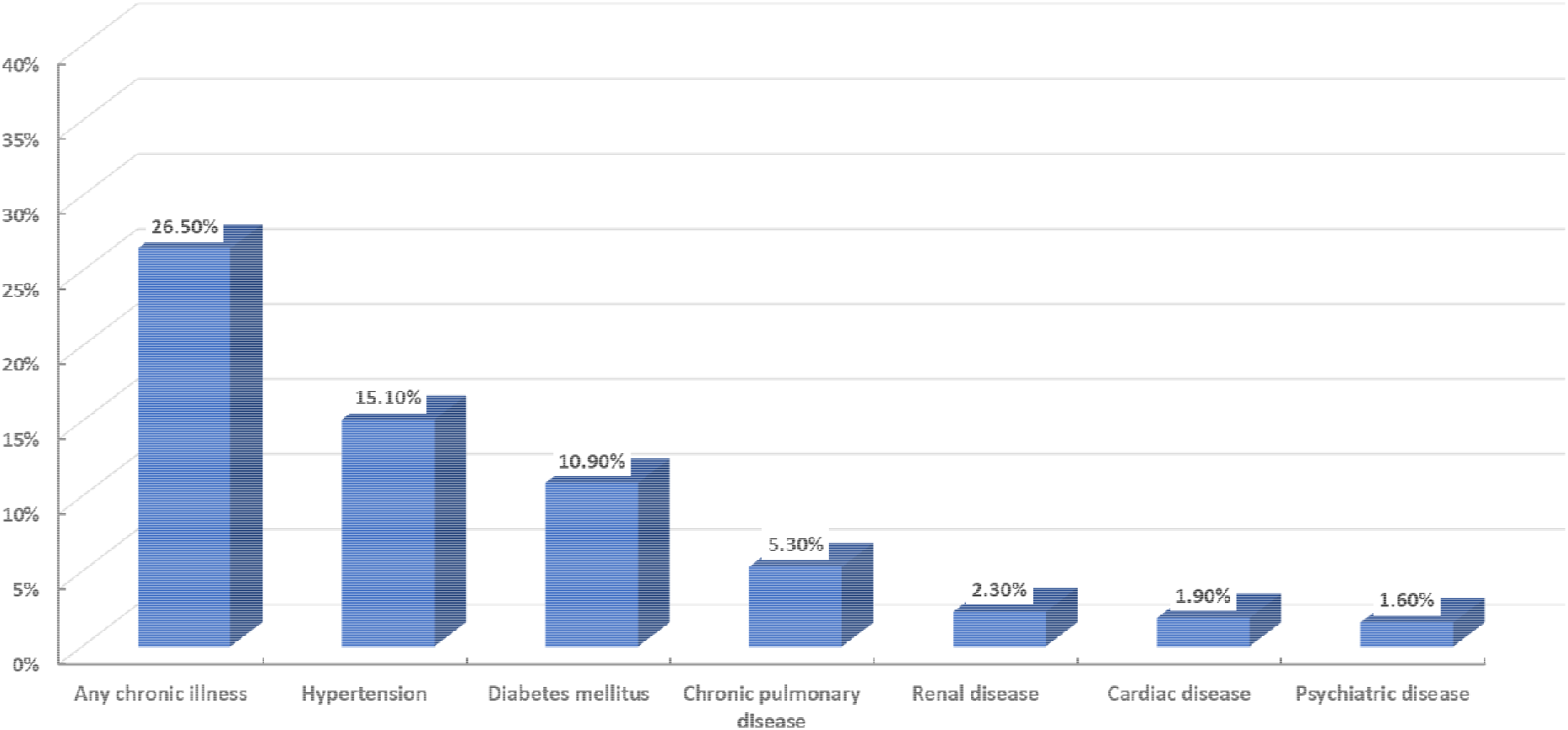
Associated co-morbidities in post-COVID-19 patients included in the study (n=430)

There was a strong positive correlation between the symptom score during the acute attack and post COVID-19 stage (P < 0.001, r=0.67) as illustrated in ***Figure (3)***.

**Figure 3.**
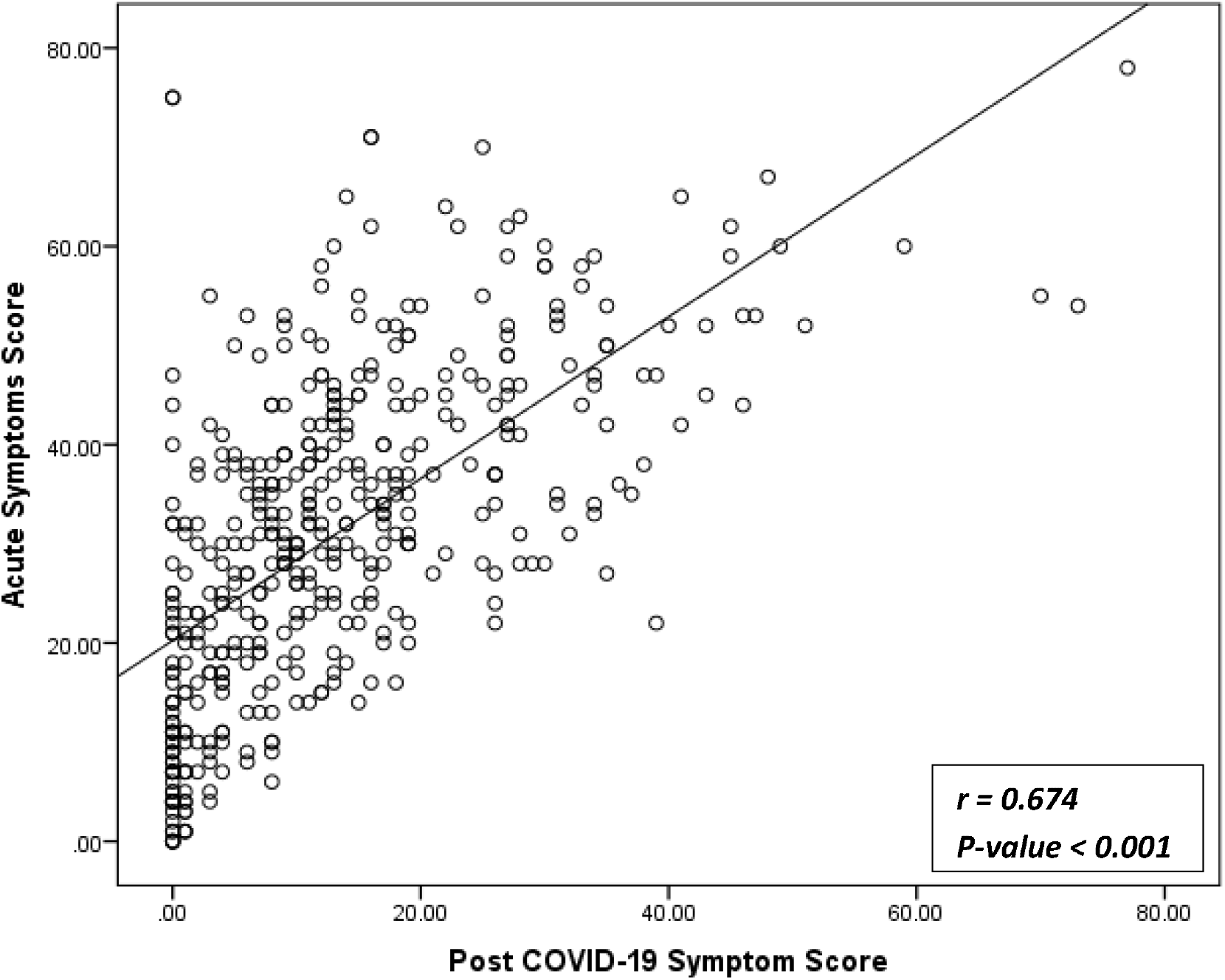
Correlation between symptoms score during acute and post covid-19 stage.

The acute phase score had 83.5% sensitivity and 73.3% specificity for the cutoff point > 18 to predict occurrence of Post-COVID symptoms ***Figure (4)***.

**Figure 4.**
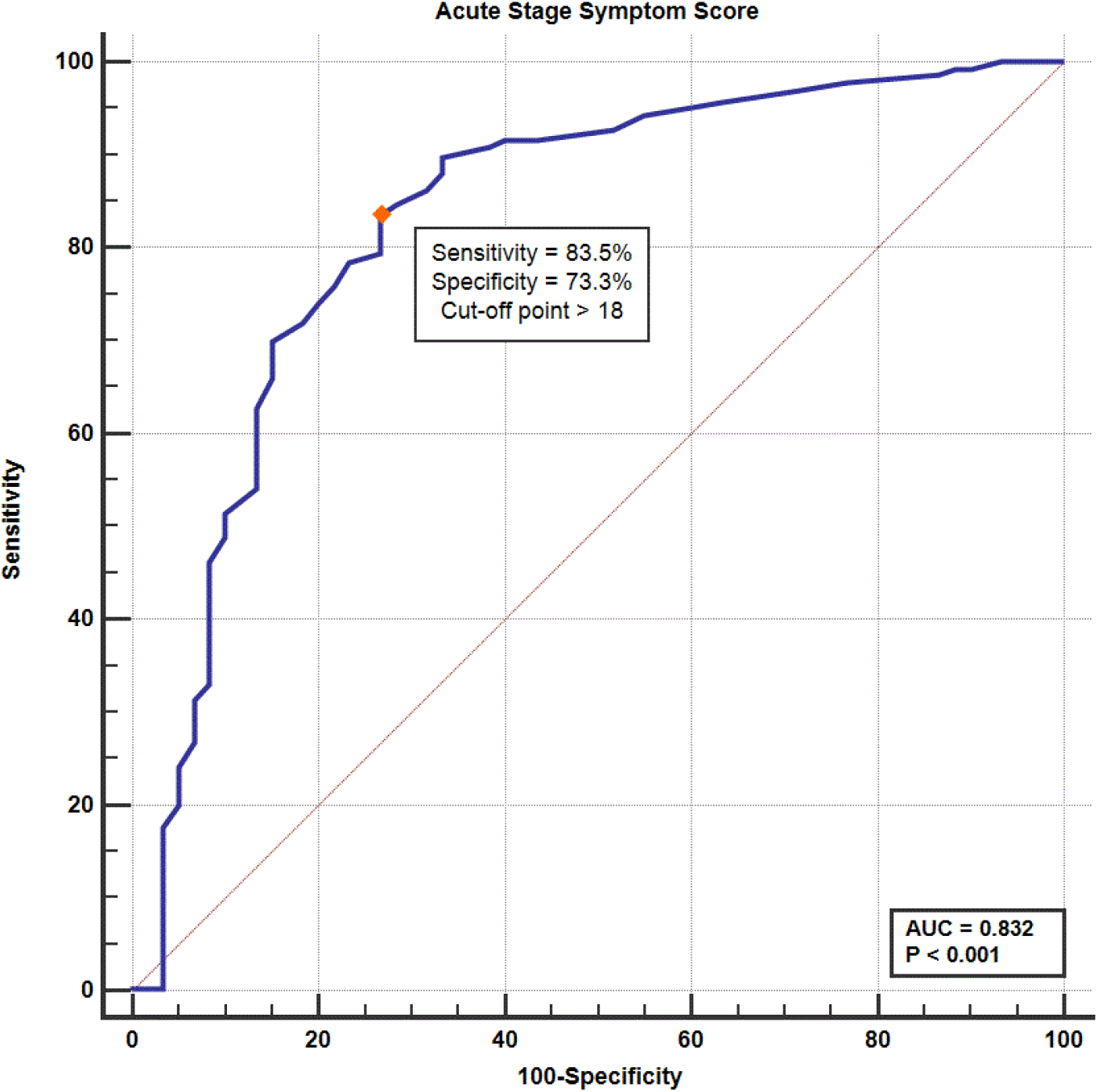
ROC curve for the acute stage symptom score as a predictor of persistent post-COVID symptoms.

## Discussion

COVID-19 is a novel illness with limited information on the post-COVID-19 symptoms. The aim of the current study was to identify the essential determinates of the different patterns of these symptoms. Most of the studied population had been still complaining of several persistent post-COVID-19 symptoms. The most frequent constitutional and neurological symptoms were myalgia (60.0%), arthralgia (57.2%), restriction of daily activities (57.0%), sleeping troubles (50.9%), nervousness and hopelessness (53.3%), while the most common respiratory and GIT symptoms were anorexia (42.6%), chest pain (32.6%), gastritis (32.3%), cough (29.3%) and dyspnea (29.1%). The mean total score of these post COVID-19 symptoms was 13.1±12.6. The main essential determinants of the persistent post COVID-19 symptoms and symptom score among the included patients were previous seasonal Influenza’s vaccination (P=0.003), and the need for oxygen therapy (P<0.001). Moreover, the most frequent pre-existing co-morbidities allied with the persistent post COVID-19 symptoms and symptom score among the study population were hypertension (P=0.039),) followed by chronic pulmonary disorders (P=0.012), and lastly the presence of any chronic disorder (P=0.004). There was a strong positive correlation between the symptom score during the acute attack and post COVID-19 stage (P<0.001, r=0.67). Furthermore, the acute phase score had 83.5% sensitivity and 73.3% specificity for the cutoff point > 18 to predict occurrence of Post-COVID symptoms.

By reviewing available literatures, about 87% of recovered from COVID-19 infection were still suffering at least one symptom one to two months after disease onset. A wide spectrum of symptoms was reported including lethargy, breathing difficulty, cough, palpitations/ tachycardia, chest pain, sleeping troubles, headache, joint ache, and deterioration of physical and mental well-being (5, 8-12).

On the other hand, an initial report of COVID-19 post-discharge complaints in China, summarized that 86.2% of cases were asymptomatic while only 9.1 had cough and 1.5% had breathing difficulty that did not affect neither daily activities nor sleep, while dizziness, headache and lethargy were not reported by those patients at all (13).

The duration of symptom resolution in included COVID-19 cases appears to be longer than that seen in community acquired pneumonia caused by bacterial pathogens. Previous studies in patients with community acquired pneumonia found that 97% of their symptoms recovered by average of ten days, while dyspnea resolved after an average of 2 weeks from the onset of the symptom, and lethargy after 3 weeks (14, 15).

It is not yet clearly recognized why some patients have persistent recovery. Long-lasting viraemia owing to vague or weak or immunological reaction(16), relapse or re-contamination (17), inflammatory and other immunological responses (18, 19), de-conditioning (20), and psychological elements such as post trauma stress syndrome may all added (21, 22). Moreover, severe COVID-19 infections necessitate management in ICU and may lead to persistent post-recovery sequelae including respiratory, physical, mental, and psychological disorders (2, 3). These complaints are stated to as post-intensive care syndrome (PICS); these sequelae can have persistent implications on the life quality (4). Patients suffering PICS commonly report higher prevalence of mental and physical disorders, which may often be long-standing (23). PICS can also cause disability and reasonable pain (24). According to Murray et al, (2020) about half percent of hospitalized patients for COVID-19 will necessitate constant care to ameliorate their long-standing consequences (6).

Post-recovery symptoms may also be predicted from the preceding corona-virus epidemics of severe acute respiratory syndrome. SARS survivors still had long-lasting lethargy, myalgia, weakness, hopelessness, psychological distress, and sleep abnormalities which may overlay with the clinical and sleep topographies of fibromyalgia and chronic fatigue syndrome (25). Myopathy due to corticosteroid use, muscle degenerative changes and weakness has similarly been described in of ARDS survivors during one-year follow-up period(26). Another study on the survivors of SARS confirmed deficits in cardio-respiratory performance in 6-minute walking test, abnormalities in the musculo-skeletal performance and quality of the life impairment (27). A similar image was described subsequently after the H1N1 influenza epidemic in 2009 (28). Following SARS, some cases suffered a decline in their mental well-being during one□year follow up period comprising nervousness, hopelessness, high incidence of posttraumatic stress disorders and psychosis (29).

Moreover, 10 years following SARS recovery, vulnerability to lung contagions, abnormality in glucose absorption and elevated levels of phosphatidylinositol persist in comparison with healthy ones (30). A recent meta□analysis found that 25% of SARS and MERS survivors had diminished lung function, quality of life and exercise capability at 6 months post-discharge (31). Similarly, the MERS convalescent cases also reported the ominously lower quality physical health for at least 14□months after the infection start, also survivors who anticipated intensive care unit admittance described an ominously minor inclusive quality of life than those with non-critical disease (32).

In the current study 29.1% of included cases had breathing difficulty. This may be explained by some persistent fibrotic changes in the lungs of COVID-19 recovered patients following the current management and discharge rules which may disturb their respiratory function (33). Moreover, patients with severe COVID-19 criteria may progress to acute respiratory distress syndrome (ARDS) and necessitate mechanical ventilation. ARDS may cause indefinite lung damage, contributing to persistent respiratory disorders after convalescence (34). Amid 33 and 75% of cases with COVID-19 necessitate mechanical ventilation, often for a long period and there are substantial long-term effects allied with prolonged period of intubation (35, 36). Those on ventilators are more susceptible to respiratory contagions, which, consecutively, making patients more vulnerable to further risk of irreversible damage of the lung tissue.

There is emergent proof advocating that pulmonary thrombo-embolism is likely an under-reported complication allied with COVID-19 that carries actually a major risk of long-standing pulmonary hypertension (37). Furthermore, prior studies have shown that acute lung injury is allied with pulmonary fibrosis on CT scans and associates with restrictive functional pattern and worse quality of life (38).

Thirty-five percent of cases included in this study had nervousness and hopelessness. Consistently, COVID-19 is linked with a major mental health problem in both the acute stage and the chronic term (39). Nervousness, hopelessness, post-traumatic stress disorder, and sleeplessness, are common behind severe corona-virus contagions (40, 41). Thirty percent of the initial 153 COVID-19 patients in the United Kingdom had psychological health troubles comprising neurosis, decline in cognitive functions, and other disorders (39). Corticosteroid therapy is also associated with the progression of psychotic complaints (42). Correspondingly, after SARS, 5%–44% complained from several mental disorders at one-year comprising nervousness, hopelessness, psychosis and greater rates of post trauma stress syndrome (29).

Myalgia (60.0%) and arthralgia (57.2%) were common complaints among patients included in this study. Correspondingly, Post-covid-19 long-lasting pain may distress patients of various age groups, but the elderly patients are the most commonly affected (43). Similarly, after the acute SARS, some patients, may evolve a Chronic Fatigue Syndrome/Myalgic Encephalomyelitis (CFS/ME) – like disease with worse sleep quality, lethargy, myalgia and hopelessness, with some incapable to coming back to their work (25).

Finally, the results of this study are challenged by some **limitations**. First, the designated sample of post-COVID-19 cases is not entirely illustrative of the all post-COVID-19 patients. Second, symptoms that initiated after the date of analysis were not verified in this survey. Third, random selection bias may be present and an inability for personal face-to-face interview in some cases. Finally, our results were made as a single point of follow-up and further follow up at 3, 6, or 12 months would aid further understanding of the progression of symptoms post-COVID-19. So, more studies and researches are desired to better appreciate, describe, and identify the persistent post-COVID symptoms in various sceneries and residents.

## Conclusions

COVID-19 is an emerging disease that can present with a diverse spectrum of long-term post COVID-19 symptoms. All patients may experience post COVID-19 symptoms, however, increased acute phase symptom severity and score above 18 together with the presence of any comorbid diseases increase the risk for persistent post COVID-19 manifestations and severity. Physicians and health care employees should aware patients, especially high-risk group, of the probable long-lasting problems of COVID-19 and reassure them to pursue medical care for any condition they may progress. Moreover, we recommend initiating multi-disciplinary post COVID-19 recovery units’ emphases on prompt and regular communication and socialization, along with physical and neurological assessments and management together with increasing scope for further detailed researches throughout those units.

## Data Availability

The datasets used and/or analysed during the current study are available from the corresponding author on reasonable request.

## List of abbreviations

DM: Diabetes mellitus
PCR: Polymerase chain reaction
BMI: Body mass index
GIT: Gastrointestinal symptoms
ROC curve: Receiver operating characteristic curve
CFS/ME: Chronic Fatigue Syndrome/Myalgic Encephalomyelitis
SARS: Severe acute respiratory syndrome
MERS: Middle East respiratory syndrome
ARDS: Acute respiratory distress syndrome
ICU: Intensive care unit
PICS: Post-intensive care syndrome

## Declarations

### Ethics approval and consent to participate

The Research Ethics Committee at the Faculty of Medicine, Aswan University, has approved the study **(IRB number: aswu /469/7/2020)** and the study was registered in Clinicaltrial.gov: NCT04479293, and all patients provided written informed consent before participation

### Consent for publication

The manuscript has been read and approved by all the authors

### Competing interests

The authors declare that they have no competing interests

### Funding

No financial support was needed

### Authors’ contributions

I G was the principal investigator, formulated the idea, wrote the first draft of discussion. AMH, collected the data, formulated the results, edited the final draft and revision, MA was responsible for methodology and statistical analysis. …. MS, HZ, MA, MM, AE, RH, NA, MS were responsible for patient’s interview and data collection….. HA, AI, KK, EF, MH were responsible for data acquisition, review search, writhing the primary draft. The manuscript has been read and approved by all the authors

## Acknowledgements

none

## Notes

### Competing Interest Statement

The authors have declared no competing interest.

### Clinical Trial

NCT04479293

### Author Declarations

aswu /469/7/2020

